# VGsim: scalable viral genealogy simulator for global pandemic

**DOI:** 10.1101/2021.04.21.21255891

**Authors:** Vladimir Shchur, Vadim Spirin, Dmitry Sirotkin, Evgeni Burovski, Nicola De Maio, Russell Corbett-Detig

**Affiliations:** HSE University, Russian Federation; European Molecular Biology Laboratory, European Bioinformatics Institute, Wellcome Genome Campus, Hinxton, Cambridgeshire, CB10 1SD, UK; Department of Biomolecular Engineering and Genomics Institute, UC Santa Cruz, California 95064

## Abstract

Accurate simulation of complex biological processes is an essential component of developing and validating new technologies and inference approaches. As an effort to help contain the COVID-19 pandemic, large numbers of SARS-CoV-2 genomes have been sequenced from most regions in the world. More than 5.5 million viral sequences are publicly available as of November 2021. Many studies estimate viral genealogies from these sequences, as these can provide valuable information about the spread of the pandemic across time and space. Additionally such data are a rich source of information about molecular evolutionary processes including natural selection, for example allowing the identification of new variants with transmissibility and immunity evasion advantages. To our knowledge, there is no framework that is both efficient and flexible enough to simulate the pandemic to approximate world-scale scenarios and generate viral genealogies of millions of samples. Here, we introduce a new fast simulator VGsim which addresses the problem of simulation genealogies under epidemiological models. The simulation process is split into two phases. During the forward run the algorithm generates a chain of population-level events reflecting the dynamics of the pandemic using an hierarchical version of the Gillespie algorithm. During the backward run a coalescent-like approach generates a tree genealogy of samples conditioning on the population-level events chain generated during the forward run. Our software can model complex population structure, epistasis and immunity escape. The code is freely available at https://github.com/Genomics-HSE/VGsim.

## 1 Introduction

The unprecedented world-wide effort to produce and share viral genomic data for the ongoing SARS-CoV-2 pandemic allows us to trace the spread and the evolution of the virus in real time, and has made apparent the need for improved computational methods to study viral evolution [1]. These data yield important insights into the effects of population structure [2–5], public health measures [6, 7], immunity escape [8,9], and complex fitness effects [10,11]. It is essential that we also have tools to accurately simulate viral evolutionary processes so that the research community can validate inference methods and develop novel insights into the effects of such complexities. However, there are no software packages capable of simulating the scale and apparent complexity of viral evolutionary dynamics during the SARS-CoV-2 pandemic.

Pandemic-scale datasets impose technical problems associated with the scalability and memory usage of computational methods. There is already substantial progress in building scalable simulators and data analysis methods for human genome data. The current state-of-the-art human genome simulator msprime [12] is capable of simulating millions of sequences with length comparable with human chromosomes. Methods such as the Positional Burrows-Wheeler Transform (PBWT) [13], its ARG-based extension tree consistent PBWT [14], and tsinfer [15] can be used to efficiently process and store genomic sequences, but all of these approaches are designed for actively recombining organisms. Moreover, the primary population models underlying these methods are the Kingman coalescent [16], the Wright-Fisher model [17, 18] and the Li-Stephens model [19]. We recently developed approaches for compressing and accessing viral genealogies that dramatically reduce space and memory requirements [20,21], but there are no viral genealogy simulation methods that can efficiently produce pandemic-scale datasets.

Coalescent models are powerful tools for studying humans, many other eukaryotes, and pathogen populations (*e*.*g*. [22]). However, their assumptions are often violated in epidemiological settings. First, the effective population size is usually modelled either as piece-wise constant or as exponential growth. However, the coalescent with exponential growth and birth-death do not result in equivalent genealogies [23]. Second, it’s not simple to use coalescent models to describe the effects of selection. If we consider the pandemic on a longer time period, basic birth-death models (e.g. [24]) are not an appropriate choice, since the reproductive rate usually decreases with time as collective immunity builds up or as the susceptible population is exhausted. These limitations are often addressed in epidemiology using compartmental models, such as SI, SIS and SIR [25], or their stochastic realisations, which are also birth-death processes.

Simulating realistic selection in backward-time models is a well-known challenging problem. A common workaround is to assume a single deterministic frequency trajectory or to generate a stochastic frequency trajectory in forward time, and then to simulate the ancestry of the samples around the selected site in a coalescent framework (e.g., [26, 27]). However, more complex models of selection, including *e*.*g*., gene-gene interactions, or epistasis, are often beyond the scope of such coalescent models. Nonetheless, epistasis is thought to be an important component of viral evolutionary processes [28, 29], and incorporating the effects of such complex evolutionary dynamics is essential for accurate simulations of evolution.

We introduce a novel simulation method that can rapidly generate pandemic-scale viral genealogies. Our approach is a forward-backward algorithm where we generate a series of stochastic events forward in time, then traverse backwards through this event series to generate the realized viral genealogy for a sample taken from the full population. Our framework includes the accumulation of immunity within host populations and of viral mutations that affect the fitness of descendant lineages. Our method is extremely fast, and can produce a phylogeny with 50 million total samples in just 88.5 seconds. The genealogies output from our simulation are compatible with phastSim [30], making it possible to generate realistic genome data for the simulated samples. This framework empowers efficient and realistic simulation of pandemic-scale viral datasets.

## 2 Design and Implementation

Our epidemiological model is a compartmental model [31] (see SI 1 for a brief introduction to compartmental models), and the realisations of the stochastic processes are drawn using the Gillespie algorithm [32]. The different compartments in our model are defined based on several interacting real-world complexities: (1) host population structure with corresponding population-specific viral frequencies and contact rates, (2) separate host infectious groups resulting from different viral haplotypes, and (3) different host susceptibility groups.

We break the simulation into two phases. In the first one (the forward pass), we generate a population-level epidemiological process which is represented as the series of events (Figure 1) resulting from the “reactions” (Table 1). These events then influence the properties of the viral genealogy, which is sampled in the second phase (the backward pass). The specific viral genealogy is sampled conditioned of the population-level epidemiological process using a coalescent framework.

**Table 1:**
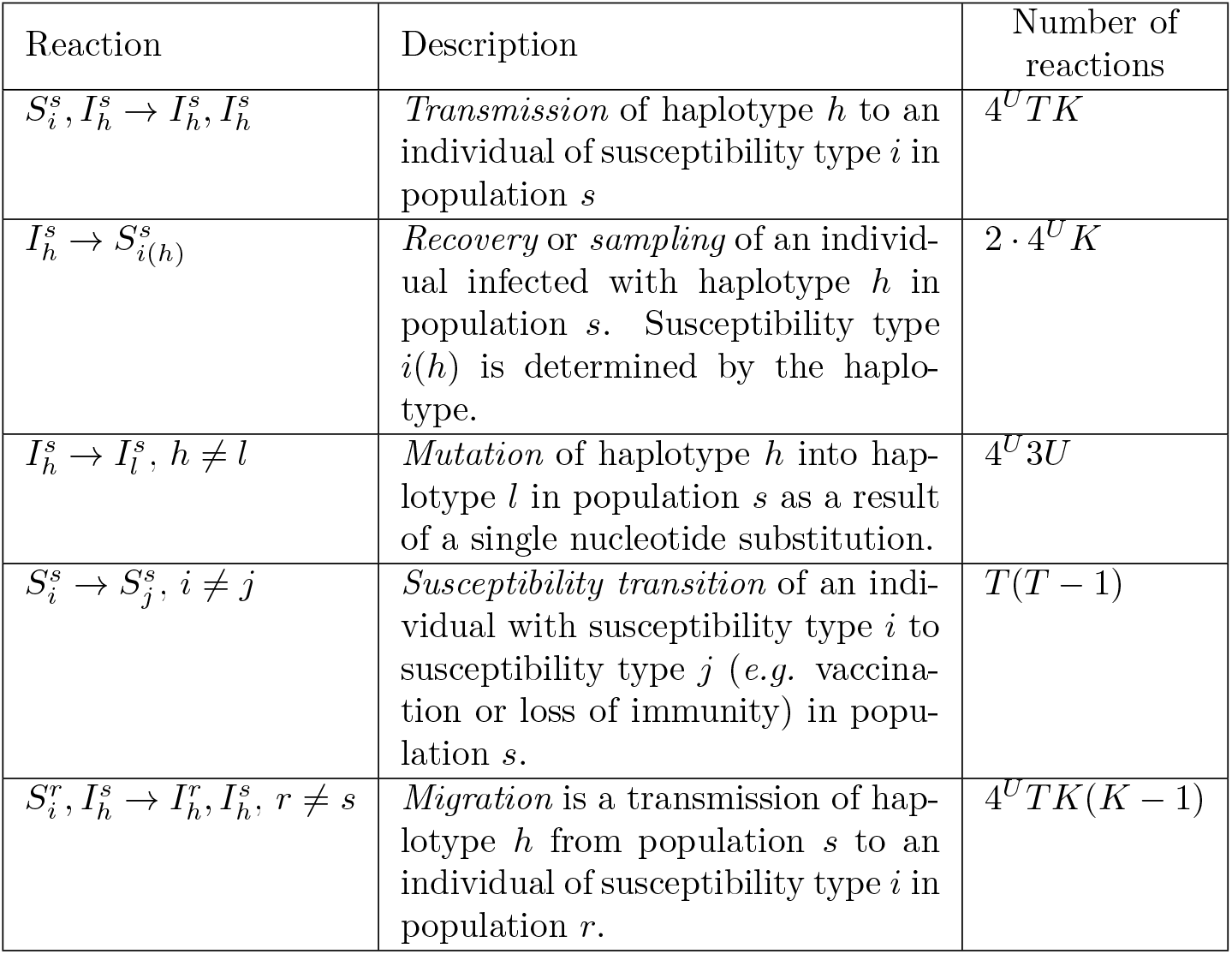
The list of reactions and corresponding epidemiological events simulated by the Gillespie algorithm in our model, and the number of reactions in each category in function of the number of mutable sites *U*, number of susceptible individuals *T*, and the number of populations *K*.

| Reaction | Description | Number of reactions |
| --- | --- | --- |
| $S_i^s, I_h^s \rightarrow I_h^s, I_h^s$ | *Transmission* of haplotype $h$ to an individual of susceptibility type $i$ in population $s$ | $4^U TK$ |
| $I_h^s \rightarrow S_{i(h)}^s$ | *Recovery* or *sampling* of an individual infected with haplotype $h$ in population $s$. Susceptibility type $i(h)$ is determined by the haplotype. | $2 \cdot 4^U K$ |
| $I_h^s \rightarrow I_l^s, h \neq l$ | *Mutation* of haplotype $h$ into haplotype $l$ in population $s$ as a result of a single nucleotide substitution. | $4^U 3U$ |
| $S_i^s \rightarrow S_j^s, i \neq j$ | *Susceptibility transition* of an individual with susceptibility type $i$ to susceptibility type $j$ (e.g. vaccination or loss of immunity) in population $s$. | $T(T-1)$ |
| $S_i^r, I_h^s \rightarrow I_h^r, I_h^s, r \neq s$ | *Migration* is a transmission of haplotype $h$ from population $s$ to an individual of susceptibility type $i$ in population $r$. | $4^U TK(K-1)$ |

**Figure 1:**
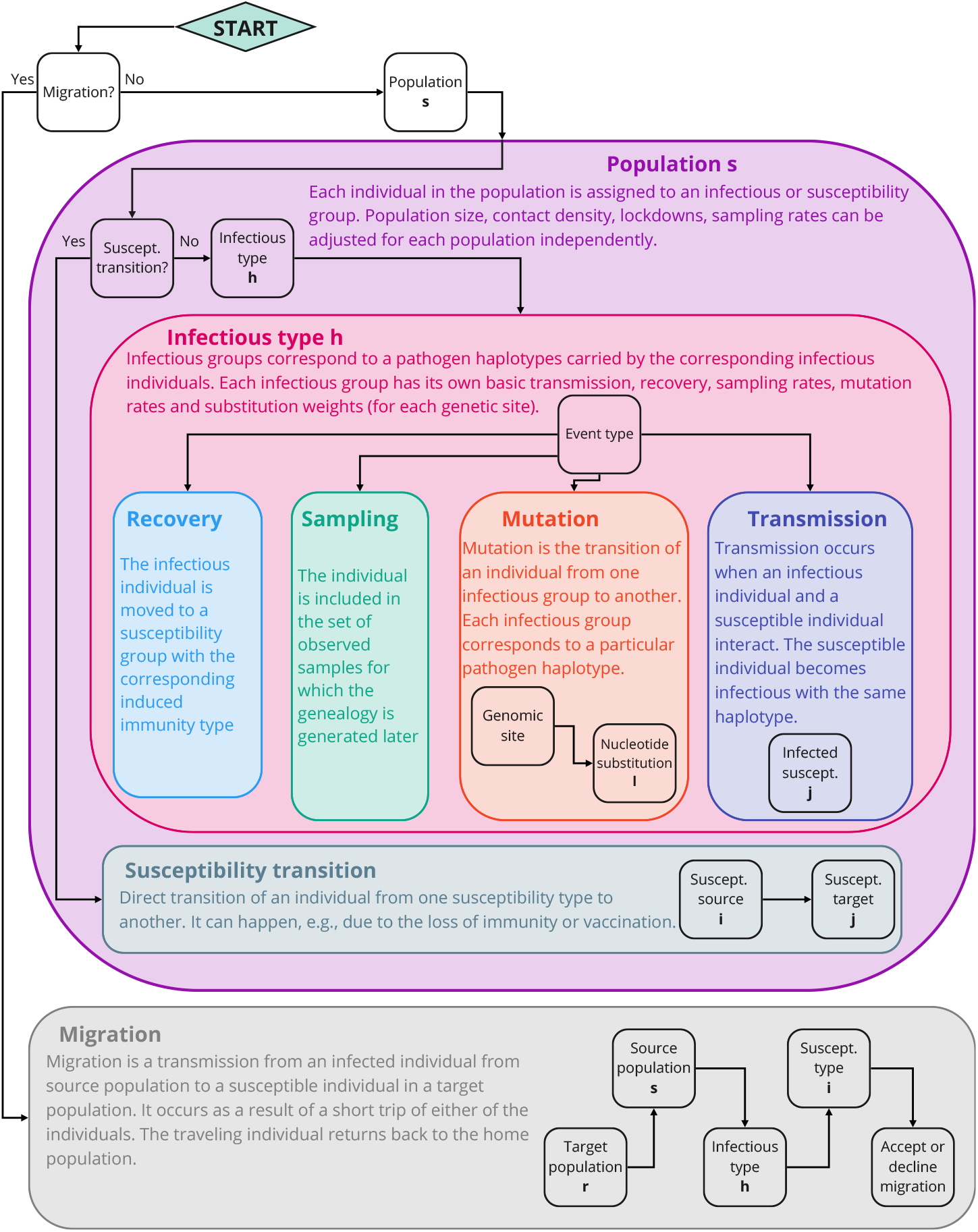
The scheme of the nested family Gillespie algorithm used to generate an event in the forward run. The corresponding reactions are listed in Table 1. Black squares correspond to the consecutive steps, where the subfamilies are chosen with the weights given by their propensities. The propensities for each step are cached and updated only if they change due to an event. For migration propensities, the rejection approach is used instead (SI 3).

Table 2 lists all the features which determine the simulation. In the beginning, the user should specify the number *U* of mutable sites (see Section 2.1), the number *T* of susceptibility types (Section 2.2), and the number *K* of populations (Section 2.3).

**Table 2:** List of features which determine the simulation scenario. All the rates are normalized by the number of individuals in a particular group (i.e. the number of individuals infected with a particular haplotype or individuals of a certain susceptibility type). The rates are measured in terms of events per time unit.

| Model | Feature | Description | Value |
| --- | --- | --- | --- |
| Epidemiological model: every parameter can be set individually for each haplotype. | Transmission rate | The expected number of new infections per time unit caused by an individual infected with haplotype $h$ if all the population were completely susceptible. | $\lambda_h \in (0; \infty)$ |
| | Recovery rate | Rate at which an infectious individual becomes recovered after being infected with haplotype $h$. | $\rho_h \in (0; \infty)$ |
| | Sampling rate | Rate at which an infectious individual is sampled after being infected with haplotype $h$. | $\zeta_h \in [0; \infty)$ |
| | Mutation rate | Rate at which a genetic site $u$ mutates. Can be set independently for each mutable site in function of haplotype $h$. | $m^u(h) \in (0; \infty)$ |
| | Substitution probabilities | The probabilities of particular nucleotide substitution at haplotype $h$ given that the mutation occurred at the site $u$. | $[p_{h_1}^u, p_{h_2}^u, p_{h_3}^u]$<br>$0 \leq p_{h_i}^u \leq 1$<br>$p_{h_1}^u + p_{h_2}^u + p_{h_3}^u = 1$ |
| | Susceptibility | The multiplier which allows to change the relative susceptibility to haplotype $h$ of an individual with susceptibility type $i$. | $\sigma_{ih} \in [0; \infty)$ |
| | Susceptibility transition rate | The rate at which susceptible individuals move from one susceptibility type to another without being infected. This allows to model the loss of immunity with time or vaccination efforts. | $[0; \infty]$ |
| Population model | Population size | Total number of individuals in population $s$. | $N^s \in (0; \infty)$ |
| | Contact density | The multiplicative modifier of transmission rate corresponding to the relative number of contacts in population $s$. It is used to describe differences in social behaviour of the host population (e.g. population density, wearing masks, lockdown effect). | $\delta^s \in [0; \infty)$ |
| | Lockdown | Conditions to impose and lift lockdown in population $k$ (determined by the proportion of infectious individuals in the population) and the contact density during the lockdown. | |
| | Sampling effort | This modifier increases or decreases the sampling rate in population $s$. | $c^s \in [0; \infty]$ |
| | Migration probability | The probability that an individual from population $s$ is temporarily visiting population $r$. | $\mu_{sr} \in [0; 1]$ |

### 2.1 Mutations

Because this simulation framework focuses on generating the viral genealogy, and not genomes, we track only mutations at genome sites that have a large positive effect on viral fitness. That is, these mutations enhance the transmissibility of the virus or lead to immunity escape. We expect this will typically be a relatively small number of mutations relative to the size of the viral genome, simplifying the problem. To efficiently model neutral genetic variation we suggest using `phastSim` [30] on a tree generated by our algorithm; the output produced by our method can be directly imported into phastSim for downstream processing.

To define the intended model of selection on new mutations, the user specifies the number *U* of mutable sites and their specific fitness effects (i.e., their effect on the birth rate). The mutations are modelled as single nucleotide substitutions, so each site has four possible variants (A, T, C and G). Mutations lead to the appearance of different haplotypes with different transmission and immunological properties. Up to *H* = 4^*U*^ different haplotypes can appear in the simulation. Each haplotype *h* can be assigned its own specific *U* mutation rates *m*^*u*^(*h*) and 3*U* substitution probabilities 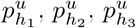, one for each site *u* and for each of the 3 possible new nucleotides at site *u*. Transmission, recovery, and sampling rates, as well as mutation rates, susceptibility, and triggered susceptibility types can be defined individually for every haplotype. Of particular interest, gene-gene, or epistatic, interactions can be flexibly modelled using this approach.

We refer to sequences carrying particular sets of variants as “haplotypes”, because two identical sequences can appear as a results of different mutation events, so they might not belong to the same clade in the final tree.

### 2.2 Epidemiological model

To model the host immunity process, we use a generalised SI-model. The compartments within each population represent different types of susceptible individuals or infectious individuals.

Different susceptible compartments in the same host population are used to model different types of immunity. These can correspond for example to host individuals that have recovered from previous exposure to different viral haplotypes. Susceptible compartments can also be used to represent different vaccination statuses. For each susceptible compartment *S*_*i*_, and for each viral haplotype *h* we consider a susceptibility coefficient *σ*_*ih*_ which multiplicatively changes the transmission (birth) rate of the corresponding haplotype. In particular, *σ*_*ih*_ = 0 corresponds to absolute resistance, similar to the R-compartment in SIR-model, but specific to individuals who have immunity type *i* and are exposed to haplotype *h*. 0 *< σ*_*ih*_ < 1 would correspond to partial immunity, while *σ*_*ih*_ *>* 1 corresponds to increased susceptibility.

Different infectious compartments within a host population correspond to individuals infected by a haplotype and can potentially infect susceptible hosts. As we mentioned in the section 2.1, the transmission rate *λ*_*h*_, recovery rate *ρ*_*h*_ and mutation rates can be set independently for each haplotype *h*. After recovery, a host individual that was infected with haplotype *h*, and therefore was in compartment 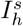 for some population *s*, is moved to the corresponding susceptibility (immunity) compartment 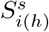. Different haplotypes might however lead to the same types of immunity.

*NB:* The evolution of individual immunity is modeled as Markovian - it is determined only by the latest infection, and does not have memory about previous infections. Whether this assumption provides an accurate approximation of the immunity dynamics within the host population is an important consideration and may depend on the specific pathogen biology. Different haplotypes can lead to the same immunity. Some immunity types can be specific, *e*.*g*., to vaccination immunity without being associated with any haplotypes at all.

The rate of transmission of viral lineages within a population also depends on how frequently two host individuals come in contact with each other. To flexibly accommodate such differences, each population *s* is assigned a contact density *δ*^*s*^ parameter. This parameter can be used to simulate differences in the local population density, social behaviours etc. The rate for an individual from susceptibility class 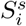 (the susceptible compartment *i* within population *s*) to be infected with haplotype *h* by another individual within population *s* is

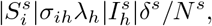

where 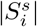 is the number of individuals in 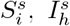 is the class of individuals infected with haplotype *h* in population *s, λ*_*h*_ is the baseline transmission rate of individuals infected with haplotype *h*, and *N*^*s*^ is the total population size of deme *s*.

Direct transitions between susceptible compartments are possible, for which users can specify a transition matrix for susceptible compartments. A transition between susceptible compartment can be used for example to model a vaccination event, or the loss of immunity with time.

### 2.3 Population model

#### 2.3.1 Demes

The population model is based on an island (demic) model. Each population is described at each point in time by its total size *N*^*s*^, number 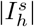 of infectious individuals with each viral haplotype *h*, number 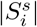 of susceptible host individuals for each susceptibility type *i*, relative contact density *δ*^*s*^, and a population-specific lockdown strategy and effectiveness.

#### 2.3.2 Lockdown

Several governments have imposed lockdowns during the COVID-19 pandemic as an effort to control the spread of SARS-CoV-2. Understanding the effects of lockdowns is a crucial concern for designing effective public health strategies. We implement lockdowns as follows. When the total number of simultaneously infectious individuals in population *s* surpasses a certain user-defined population-specific percentage (e.g. 1%) of the population size *N*^*s*^, the lockdown is imposed and the contact density *δ*^*s*^ is changed to a new (typically lower) lockdown- and population-specific value. When the percentage of the infectious individuals drops below a user-specified value (e.g. 0.1%) the lockdown is lifted and the contact density in population *s* reverts to its initial value *δ*^*s*^.

#### 2.3.3 Migration

Migration is described by a matrix *µ*_*sr*_ which defines the probabilities at which an individual from source population *s* can be found in target population *r*. In our model, new infections occur from the contact between infectious individuals from one population and susceptible individuals from the second population. It can be due to the travel of a susceptible individual to a source population, where it contracts an infection, and then returns back to the home population (first term in equation 1); or, to the travel of an infectious individual into a target population where this individual transmits the infection to a susceptible individual (second term in equation 1). The derivation of each term is similar to the derivation of within-population transmission (see SI equation 1). This model corresponds to short-term travel such as tourist or business trips, where an individual returns soon back to the home population. The proposed process is different from the traditional migration modelling in the population genetics, when an individual moves to a new population and remains there. The rate at which new infections of individuals with immunity *i* in population *r* are caused by haplotype *h* in population *s* is

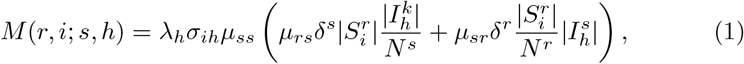

where *µ*_*ss*_ = 1− ∑ *q*≠*s µ*_*sq*_ is the probability that an individual originally from population *s* is currently not in a different population. Since it is computationally demanding to keep track of how each migration rate between each pairs of compartment is affected by each simulation event, instead we keep track of cumulative upper bounds on such migration rates (see SI 3 for details). In the case a potential migration event is sampled according to these upper bounds, we then proceed to calculate the precise migration rates and only sample a specific migration event according to its own exact rate. This is efficient when cross-population transmissions (migrations) are rare compared to within-population transmissions. This algorithmic implementation might perform suboptimally if population structure is extremely weak.

### 2.4 Sampling

Sampling is modelled using a continuous sampling scheme. In this scheme every individual infected with haplotype *h* in population *s* is sampled at rate *c*^*s*^*ζ*_*h*_, the product of the haplotype-specific sampling rate *ζ*_*h*_ and the population modifier *c*^*s*^. Sampled individuals then instantly recover and are moved to susceptible group 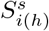, effectively increasing the recovery rate *ρ*_*h*_ for 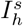 by *c*^*s*^*ζ*_*h*_. Alternatively, one can think about this sampling scheme as setting the recovery rate for 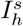 to *ρ*_*h*_ + *c*^*s*^*ζ*_*h*_ and sampling an individual in 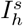 upon its recovery with probability *c*^*s*^*ζ*_*h*_*/*(*ρ*_*h*_ +*c*^*s*^*ζ*_*h*_). More details can be found in supplementary information 7.

### 2.5 Algorithm

The simulation process is split into two parts, forward and backward. In the forward run, a chain of events (including sampled cases) describing the dynamics of the epidemiological process at the population level is generated with the Gillespie algorithm [32]. In the backward run, our method simulates the genealogy of the samples in a coalescent-like manner while conditioning on the events generated during the forward run.

#### 2.5.1 Forward run

The forward run generates a chain of events which reflects the dynamics of the pandemic. Our implementation of the Gillespie algorithm is based on three algorithmic ideas: logarithmic direct method [32] (the events, or “reactions”, are organised in nested families, Figure 1), rejection-based approach [33] for migrations (see SI 3 for details), and organising propensity dependencies to avoid updating those propensities which are not affected by events [34]. Details are given in SI 4.

#### 2.5.2 Backward run

The backward run randomly builds a genealogical tree of the samples while conditioning on this chain of events generated in the forward run.

All of the ancestral lineages of the samples generated in the forward run belong to one of the infectious compartment corresponding to a haplotype *h* in a specific population *s*. Lineages are exchangeable within each compartment. Conditional on any event generated in the forward run, it is straightforward to calculate the probability that the event affected zero, one or two sample ancestral lineages in the backward run (see SI 5 for details).

#### 2.5.3 Implementation details

`VGsim` provides a convenient Python user interface. Performance-critical parts are implemented in C++ via Cython [35]. The dependencies are kept to a minimum: NumPy [36] and `mc lib` —a small wrapper of the NumPy C API for generation of pseudorandom numbers in Cython [37].

## 3 Results

### 3.1 Forward run performance

To test the scalability of the population model, we performed simulations with *K* = 2, 5, 10, 20, 50 and 100 total host populations with 2 10^9^*/K* individuals in each and generated 10 million events (see Figure 1) in each run (see Table 3). There are 16 haplotypes resulting from two segregating sites with mutation rates 0.01 in each of them (this is unrealistically high, but it ensures that all the haplotypes appear in the simulation), and three susceptibility group with the first group corresponding to the absence of immunity, the second group corresponding to partial immunity and the last one corresponding to resistance to all strains. The transmission rate is *λ* = 2.5 for all haplotypes except one, and *λ* = 4.0 for this last haplotype. The recovery rate is *ρ* = 0.9, the sampling rate is *ζ* = 0.1 (so, the effective reproductive number is 2.5 which approximately correspond to SARS-CoV-2 [38] if the time unit is interpreted as one week). All the migration probabilities were set to *µ* = *M/*(*K*−1), where *M* is the cumulative migration rate from a population. The runtime of the forward algorithm does not depend only on the cumulative migration rate *M*, but also on the percentage of potential migrations rejected by the algorithm (see Section 2.3.3 for details), which appears to grow with *M*. However, the effect on runtime is relatively modest (in contrast to the naive algorithm which is quadratic in the number of populations, see Table 1) indicating that this approach scales well to pandemic-scale simulations.

**Table 3:** Run time to generate 10 million events. The second number is the percentage of discarded events (due to migration acceptance/rejection). There are 16 = 4^2^ haplotypes and 3 susceptible compartments. The sampling rate is set to *ζ* = 0.1, recovery rate is *ρ* = 0.9, transmission rate is *λ* = 2.5. The tests were run on a server node with Intel Xeon Gold 6152 2.1-3.7 GHz processor and 1536GB of memory.

| Cumulative<br>migration<br>probability $M$ | Number of demes $K$ | | | | | |
| --- | --- | --- | --- | --- | --- | --- |
|  | 2 | 5 | 10 | 20 | 50 | 100 |
| 0.001 | 28.7s | 30.0s | 31.9s | 35.1s | 47.2s | 69.3s |
|  | 0.09% | 0.12% | 0.11% | 0.11% | 0.11% | 0.12% |
| 0.002 | 29.2s | 30.4s | 32.3s | 35.3s | 47.0s | 70.1s |
|  | 0.17% | 0.21% | 0.16% | 0.2% | 0.21% | 0.2% |
| 0.005 | 29.4s | 30.7s | 32.5s | 35.6s | 48.1s | 70.4s |
|  | 0.33% | 0.51% | 0.25% | 0.46% | 0.52% | 0.43% |
| 0.01 | 29.4s | 30.6s | 32.9s | 35.5s | 48.0s | 70.9s |
|  | 0.75% | 1.16% | 0.85% | 0.78% | 0.93% | 0.95% |
| 0.1 | 30.3s | 31.9s | 33.8s | 37.0s | 50.3s | 73.0s |
|  | 2.04% | 6.56% | 6.15% | 6.94% | 5.98% | 5.08% |

### 3.2 Backward run and overall performance

Our implementation of the backward run algorithm relies on an efficient and compact tree representation, a Prüfer-like code [39]. Each node is associated with an index in an array, and the corresponding entry in the array is the index of the parent node. The time needed to generate a tree mainly depends on two factors: the total number of events generated in the forward run, and the total number of samples in a tree. We report the execution time of the backward run in Table 4. The combined approach is sufficiently fast that it can be used to generate many replicate simulations as is often required to validate empirical methods and to train model parameters. Table 4 also shows the forward time, the total number of generated events and the total number of infected individuals over the simulation for various sampling rate (where the sampling rate *ζ* = 0.01 is 1 in 100 cases is sampled, *ζ* = 0.1 corresponds to 1 in 10 cases is sampled, and *ζ* = 1.0 means that every case is sampled), and various sample sizes. The simulation assumes the absence of immunity after infection (SIS-model), which allows to run the simulation sufficiently long to collect enough samples (instead, with an SIR-model susceptible individuals can be exhausted before the desired number of samples is simulated).

**Table 4:** Run time in seconds to generate a random genealogy for a sample of a certain size for different sampling rates. The execution time is shown split into the time demand for the forward run and the one for the backward run only. We simulated 16 = 4^2^ haplotypes and no host immunity. The recovery rate is *ρ* = 1.0 −*ζ*, with *ζ* the sampling rate, while the transmission rate is *λ* = 2.5 for all 16 haplotypes. The tests were run on a server node with an Intel Xeon Gold 6152 2.1-3.7 GHz processor and 1536GB of memory.

| Sampling rate |  | Sample size<br>(number of tree leaves) |  |  |  |  |  |
| --- | --- | --- | --- | --- | --- | --- | --- |
| | | $10^5$ | $10^6$ | $5 \cdot 10^6$ | $10^7$ | $5 \cdot 10^7$ | $1.5 \cdot 10^8$ |
| 0.01 | Forward time | 27.84s | 290.86s<br>(4min 50.86s) | 1275.53s<br>(21min 15.53s) | 2487.73s<br>(41min 27.73s) | 11295.01s<br>(3h 8m 15.01s) | 34558.86s<br>(9h 35m 58.86s) |
|  | Backward time | 0.85s | 7.44s | 26.93s | 50.27s | 217.51s<br>(3min 37.51s) | 813.25s<br>(13min 33.25s) |
|  | Memory | 1.67MB | 10.87GB | 49.54GB | 94.64GB | 442.69GB | 1.34TB |
|  | Total number of generated events | 34,038,092 | 286,381,088 | 1,120,365,070 | 2,121,897,004 | 9,878,131,708 | 30,152,423,891 |
|  | Total number of infections | 24,040,769 | 185,954,943 | 619,559,504 | 1,119,957,985 | 4,994,200,627 | 15,121,211,248 |
| 0.1 | Forward time | 2.18s | 29.89s | 154.15s<br>(2min 34.15s) | 296.43s<br>(4min 56.43s) | 1283.01s<br>(21min 23.01s) | 3470.47s<br>(57min 50.47s) |
|  | Backward time | 0.1s | 0.96s | 4.68s | 8.99s | 34.2s | 90.29s<br>(1min 30.29s) |
|  | Memory | 1.68MB | 1.68MB | 5.51GB | 12.5GB | 53.27GB | 143.32GB |
|  | Total number of generated events | 3,491,562 | 34,125,248 | 155,922,768 | 285,874,161 | 1,120,657,092 | 3,122,658,422 |
|  | Total number of infections | 2,489,943 | 24,101,573 | 105,814,516 | 185,656,462 | 619,705,716 | 1,619,831,406 |
| 1.0 | Forward time | 0.23s | 2.2s | 13.63s | 30.32s | 154.99s<br>(2min 34.99s) | 405.39s<br>(6min 45.39s) |
|  | Backward time | 0.01s | 0.15s | 0.92s | 2.08s | 11.35s | 32.48s |
|  | Memory | 1.67MB | 1.68MB | 1.66MB | 1.67MB | 5.54GB | 20.9GB |
|  | Total number of generated events | 350,517 | 3,492,789 | 17,271,140 | 34,113,125 | 155,899,482 | 401,912,500 |
|  | Total number of infections | 250,290 | 2,490,805 | 12,261,217 | 24,093,104 | 105,799,613 | 251,613,148 |

To showcase the limit of applicability of our simulator, we also show in Table 4 the computational demand for the simulation of an unrealistically large (for now) genealogy of 150 million samples (with 1 in 100 cases sampled), for which we almost reached the memory limit available on our supercomputer node (1536GB) [40]. The total number of infections in the population is more than 15 billion cases, with the total number of events being more than 30 billions. The forward run time was approximately 9.5 hours and the backward run time was 13.5 minutes.

### 3.3 Comparison with other simulators

There are many epidemiological simulators which are capable of producing viral genealogies. Agent-based simulators (e.g. `nosoi` [41], `FAVITES` [42]) allow to create very detailed models, because every agent’s parameters can be set individually. The trade-off is they are computationally demanding, so only relatively small scenarios can be modelled. Other simulators (e.g. `MASTER` [43] and `TiPS` [44]) implement Gillespie algorithm for compartmental models, but they currently lack a simple user interface, instead requiring users to specify the full set of reaction equations, and they might be not specifically optimised for epidemiological purposes. On the other hand, both `MASTER` and `TiPS` implement approximate methods (tau-leaping and hybrid approaches), which decrease the time of forward simulation by orders of magnitude and hence might outperform our simulator depending on simulation scenario. `VGsim` is optimised to scale for large epidemics and genealogies, though approximate approaches are not available in the current implementation. It also has a simple and flexible user interface which helps merge together several complexities (epidemiology, evolution, population structure and cross-immunity). The detailed discussion of different simulating frameworks and detailed comparisons with them can be found in SI 8.

### 3.4 Simulating realistic nucleotide mutations

Our simulation framework generates a phylogenetic tree, and if the user specifies a scenario with strongly selected mutations, these are included in the output; we, however, do not include a method for simulating many neutral variants. To facilitate studies that require full viral genome sequences we have made the output of our approach compatible with that of `phastSim` [30]. Briefly, a user can easily load the output of our software into `phastSim`, and `phastSim` will generate neutral mutations, while leaving previously determined selected mutations unaffected.

## 4 Availability and Future Directions

`VGsim` is freely available from https://github.com/Genomics-HSE/VGsim under GPL-3.0 License. It is tested for Python 3.6 and later under Ubuntu and macOS. The documentation and tutorials are published at https://vg-sim.readthedocs.io/.

The future development of `VGsim` will include the following updates. We will consider improving performance by adding the *τ* -leaping algorithm and optimizing memory usage to handle larger numbers of genetic sites. We will also extend the available models by adding super-spreading events, life-cycle compartments, and new sampling schemes. We will also add recombination events, though they seem to be relatively rare [45] and so far are not a major driver of SARS-CoV-2 genetic diversity and evolution.

`VGsim` is particularly useful for simulating large datasets, in particular, in those cases when agent-based simulators become inefficient (see SI for more detailed discussion 8). It is primarily optimised for the studies of world-wide pandemic scenarios, and it is motivated by the features of the ongoing SARS-CoV-2 pandemic. We plan for the future to add more features which would generalise its applicability to different pathogens (e.g. with complex life-cycle). Further possible optimisations of our algorithm will also be investigated.

Our implementation allows simulations of scenarios with a few loci with strong phenotypic effects. However, we cannot simulate the effect of many loci with mild fitness effects. While mild and widespread fitness effects can be simulated by phastSim, they are simulated in a typical phylogenetic way (using a substitution codon matrix with specifiable nonsynonymous/synonymous ratios) and so their impact on the tree shape and epidemiological dynamics are neglected.

## 5 Conclusion

We developed a fast simulator `VGsim` which can be used to produce genealogies of millions of samples from world-scale pandemic scenarios. Our method models many major aspects of epidemiological dynamics: viral molecular evolution, host population structure, host immunity, vaccinations and lockdowns. We expect that `VGsim` will be a useful tool in method validation and in simulation-based statistical inference.

The performance of our simulator should meet the performance requirements of most studies. The flexible Python API, combination of epidemiological (including cross-immunity), population and evolutionary models make it a timely tool for the modern and future research.

## Data Availability

There is no data and reagent used in the paper. The code is available at the GitHub repository associated with this project: https://github.com/Genomics-HSE/VGsim.

## 6 Acknowledgment

We are thankful to Mikhail Shishkin for testing VGsim on the Apple Silicon M1 processor. VSh, VSp, DS, RCD were funded within the framework of the HSE University Basic Research Program. EB acknowledges support within the Project Teams framework of MIEM HSE. VSh was supported by RFBR grant 20-04-60556 while working on section 2.3. This research was supported in part through computational resources of HPC facilities at NRU HSE. RCD was supported in part by NIH/NIGMS R35GM128932. NDM was supported by the European Molecular Biology Laboratory.

