## Supplementary information for "VGsim: scalable viral genealogy simulator for global pandemic"

November 2021

#### 1 Introduction to compartmental models

To get some basic intuition behind the compartmental models used in the forward pass, consider the simple case of a single population, a single viral haplotype, and an SIS compartmental model. In this model every individual can be either susceptible (S) or infectious (I). If a susceptible individual meets an infectious individual, there is a chance $x$ that the infectious individual infects the susceptible one. All individuals have the same contact rate $y$ (the number of other individuals they meet in a time unit). Assume there are $|S(t)|$ and $|I(t)|$ susceptible and infectious individuals at time $t$, and the constant total population size is $N$. For a single susceptible individual the chance that the next random encounter with another person would be with an infectious individual is $|I(t)|/(N-1)$ (though $N$ is usually large, so $N-1 \approx N$). So the rate at which a single susceptible individual becomes infected is $\lambda' = \lambda\frac{|I(t)|}{N}xy$ (contact rate times the probability that the second individual is infectious times the probability of transmission). The total rate of a new infection in the population will be $\tilde{\lambda} = |S(t)|\lambda'$. Similarly, infectious individuals recover with some rate $\rho$, and the total recovery rate in the population is $\tilde{\rho} = \rho|I(t)|$. From the theory of Poisson processes, it follows that the total rate of events in the population is simply the sum $\tilde{\lambda} + \tilde{\rho}$, and the waiting time of the first event is distributed exponentially

with rate (parameter) $\tilde{\lambda} + \tilde{\rho}$, and with probability $\tilde{\lambda}/(\tilde{\lambda} + \tilde{\rho})$ the next event is an infection. This gives a straightforward method to simulate continuous-time epidemiological dynamics under this model. This approach is easily generalised to an arbitrary number of compartments. The SIS model is the basis for all the compartmental models we implemented, and in particular we expand on the basic model (as described below in details) by allowing different types of S and I compartments in the same simulation.

### 2 Migration

In our model, migrations are interpreted as visits of individuals from one population into another. The expected number of individuals present at any time in population $s$ is

$$N_f^s = \sum_r \mu_{rs} N^r,$$

where $N^r$ is the number of individuals native to population $r$, $\mu_{rs}$ is the probability to find an individual native of population $r$ in population $s$ (notice that $\mu_{ss} = 1 - \sum_{r \neq s} \mu_{sr}$). This model is accurate in the case of large population sizes. For small populations, agent-based simulators are more appropriate (see SI 8).

The rate of transmission of haplotype $h$ to an individual with immunity $i$ within population $s$ is

$$\lambda_h \sigma_{ih} \delta^s \left(\mu_{ss} |I_h^s|\right) \frac{\mu_{ss} |S_i^s|}{N_f^s} = \lambda_h \sigma_{ih} \delta^s \mu_{ss}^2 \frac{|I_h^s||S_i^s|}{N_f^s}, \tag{1}$$

where, we remind, $\lambda_h$ is the base transmission rate of the haplotype $h$, $\sigma_{ih}$ is the susceptibility of immune group $i$ to haplotype $h$, $\delta^s$ is the contact density within population $s$, and $\mu_{ss}$ is the probability that an individual native to population $s$ is in their home population at the time of the event.

### 3 Efficient algorithm for migrations

Instead of updating every particular propensity for infectious and susceptible compartments from different populations, we rely on an upper-bound for propensities corresponding to migration events. If a potential migration is sampled, the algorithm then calculates the actual propensity and accepts or rejects the migration event with probability equal to the ratio of the actual propensity and its upper bound.

Here we derive the upper bounds on migration rates. For this purpose, we set $\Sigma_h = \max_i \sigma_{ih}$, $\Lambda = \max_h \lambda_h \Sigma_h$. Then the following holds

$$\sum_{i,h} M(r,i;s,h) \leq \Lambda \sum_{i,h} \left( \frac{\mu_{rs}\delta^s}{N_f^s} + \frac{\mu_{sr}\delta^r}{N_f^r} \right) |I_h^s||S_i^r|$$
$$= \Lambda \left( \frac{\mu_{rs}\delta^s}{N_f^s} + \frac{\mu_{sr}\delta^r}{N_f^r} \right) S^r I^s,$$

where $M(r,i;s,h)$ is defined by equation 1, $S^r = \sum_i |S_i^r|$ and $I^s = \sum_h |I_h^s|$. Denote $M_{sr} = \frac{\mu_{rs}\delta^s}{N_f^s} + \frac{\mu_{sr}\delta^r}{N_f^r}$. So, the total migration (or introduction) rate from source population $s$ into target population $r$ is given the upper bound of $\Lambda M_{sr} S^r I^s$.

We finally get the upper bound for the total migration rate:

$$\sum_{s,r} \Lambda M_{sr} S^r I^s \leq \Lambda \left( \max_{s,r:s\neq r} M_{sr} \right) S_g I_g,$$

where $S_g$ and $I_g$ are the total number (over all populations) of susceptible and infectious individuals in the simulation.

If the algorithm samples a potential migration, it samples $s, r, h$ and $i$ with the probabilities $\frac{|S_i^r||I_h^s|}{S_g I_g}$, and accepts the migration with probability $\frac{\lambda_h \sigma_{ih}}{\Lambda \max_{s,r} M_{sr}}$. Otherwise, the migration is discarded and the algorithm proceeds to the next iteration. The logical basis of this approach follows from the additivity of Poisson processes (similarly to the reasoning behind the standard Gillespie algorithm [1]).

### 4 Forward run

The forward run consists of sampling events which determine the development of the epidemiological process at the population level. In order to efficiently sample events using the Gillespie algorithm, one needs to iteratively 1) sample an event given the reaction propensities, 2) update the reaction propensities.

Event sampling is organised similarly to the logarithmic direct method [1] which groups all the reactions in subsets which can be represented as a tree. We split the reactions by the following logic represented in Figure 1

- First, the algorithm chooses between a migration event or a within-population event.
- For migration events the following parameters are chosen subsequently: target population (where the new introduction occurs), source population of the infection, introduced haplotype, immunity type of the individual contracting the infection, rejection or acceptance of the migration event (see below).
- All other events are considered to be within-population; for these, first the population of the event is sampled.

- Then, choose if the event is a susceptibility transition.
- For susceptibility transitions, choose the source and target susceptibility groups.
- Otherwise, choose the infectious compartment (haplotype) involved in the event.
- Finally, choose the type of event: recovery, sampling, mutation or transmission.
- For mutations: the genomic site and the nucleotide substitution are chosen subsequently.
- For transmissions: the immunity type of the individual being infected is chosen.

As we explained in SI 3, instead of keeping propensities for each infectious-susceptible interaction between populations, we keep track of their upper bound and apply a rejection-based algorithm [2] to the sampling of migration events.

We cache the total propensities of each population at each step of the algorithm. We also keep track of the propensity dependencies, similarly to the optimised direct algorithm [3], to avoid updating propensities which are not affected by the reactions.

### 5 Backward run

The backward run is carried out backward in time, starting from the last sampling event simulated during the forward run. $|I_h^s|$ is again the total number of individuals carrying haplotype $h$ in population $s$ at a given time. Denote by $\mathbb{L}_h^s$ the set of sample ancestral lineages with haplotype $h$ in population $s$ at the considered time, and define $L_h^s = |\mathbb{L}_h^s|$ its size.

A new infection forward in time corresponds to a coalescence between two lineages backward in time. Becoming non-infectious the forward in time corresponds to a recovered individual becomes infectious backward in time. Mutations simply revert from the derived allele back to ancestral allele when considered backward in time. Migration-associated transmission correspond backward in time to coalescent events between lineages in different populations.

A more detailed list of possible events for the backward run and their rates is the following:

- New infection with haplotype $h$ in population $s$. Two sample ancestral lineages coalesce with probability

$$\frac{\binom{L_h^s}{2}}{\binom{|I_h^s|}{2}},$$

which is the ratio of the number of pairs of lineages over the number of pairs of infected individuals. If indeed a coalescent event is sampled, we randomly choose and remove a pair of lineages $l_1, l_2$ from $\mathbb{L}_h^s$, add an ancestral node $l_a$ into the sample genealogy, and set the parent of $l_1$ and $l_2$ to $l_a$. In any case $|I_h^s| \leftarrow |I_h^s| - 1$.

- Recovery of an individual infected with haplotype $h$ in population $s$. Becoming non-infectious corresponds to a recovered individual becoming infectious backward in time. Hence, $|I_h^s| \leftarrow |I_h^s| + 1$.

- Sampling an individual infected with haplotype $h$ in population $s$. As with recovery, $|I_h^s| \leftarrow |I_h^s| + 1$. Also, a new leaf node is added to the genealogy, and the corresponding lineage is added into the set $\mathbb{L}_h^s$.

- Mutation transforming haplotype $h$ into derived haplotype $m$ in population $s$. This mutation happens to a sample ancestral lineage with probability $L_m^s/|I_m^s|$. In this case we randomly choose a lineage from $L_m^s$ and move it into $L_h^s$. In any case update

$$|I_m^s| \leftarrow |I_m^s| - 1$$
$$|I_h^s| \leftarrow |I_h^s| + 1.$$

- Transition between susceptible compartments (immunity change). This does not have any immediate effect on the genealogy.

- Migration of haplotype $h$ from source population $s$ into target population $r$. First, with probability $L_h^r/|I_h^r|$ a sample ancestral lineage is affected by the migration. In this case we randomly choose a lineage from $\mathbb{L}_h^r$ and move it into $\mathbb{L}_h^s$. With probability $(L_h^s - 1)/|I_h^s|$ this lineage coalesces with a sample ancestral lineage in the source population. In this case we randomly draw a lineage from $\mathbb{L}_h^s$ (except for the one which was just moved there), and update the genealogy similarly to the case of a new infection. In any case update

$$|I_h^r| \leftarrow |I_h^r| - 1$$

### 6 VGsim's Python API

VGsim's API allows initializing such a model with thousands of reactions in a few lines of Python code. With $U = 2$ mutable sites, $K = 100$ populations and $T = 3$ susceptibility types, there are more than $5 \cdot 10^5$ reactions.

```
import VGsim
number_of_sites = 2
populations_number = 100
number_of_susceptible_groups = 3
simulator = VGsim.Simulator(number_of_sites, populations_number,
    number_of_susceptible_groups, seed=1234)
```

Listing 1: VGsim initialisation

Now we set the transmission ($\lambda = 2.5$), recovery ($\rho = 0.99$), sampling ($\zeta = 0.01$) and mutation ($m = 0.00002$) rates, and the susceptibility type ($i = 1$, enumeration starts from 0) after recovery for all the haplotypes. These rates correspond to SARS-CoV-2 wild-type parameters if time measured in a unit of one week.

```
simulator.set_transmission_rate(2.5)
simulator.set_recovery_rate(0.99)
simulator.set_sampling_rate(0.01)
simulator.set_mutation_rate(0.00002)
simulator.set_susceptibility_type(1)
```

Listing 2: Setting baseline rates.

Then the parameters of this model can be fine-tuned (from now on all the parameters might be non-realistic and used only to demonstrate different features of the API). In the next few lines we set the following features: a transmission rate $\lambda = 4.0$ of all haplotypes carrying allele $G$, migration probabilities $\mu_{sr} = 10/365$ assuming every individual spends on average 10 days outside their population of origin, vaccination efforts corresponding to susceptibility transitions from types 0 and 1 into type 2.

```
simulator.set_transmission_rate(4.0, haplotype='G*')
simulator.set_migration_probability(total_probability=10/365)
simulator.set_immunity_transition(0.35, target=2)
```

Listing 3: Adjusting the simulated model.

Finally we tune the susceptibility. There will be partial immunity $\sigma_1 = 0.2$ after recovery to all haplotypes except for 'GT'. Previous infection increases the susceptibility to haplotype 'GT' by 10%: $\sigma_{i,GT} = 1.1$. Vaccine gives complete resistance to all haplotypes. Finally, run the simulation for 10 time units and no more than $10^7$ events.

```
simulator.set_susceptibility(0.2, susceptibility_type=1)
simulator.set_susceptibility(1.1, susceptibility_type=1, haplotype=
    'GT')
simulator.set_susceptibility(0.0, susceptibility_type=2)
simulator.simulate(10000000, time=10)
```

Listing 4: Setting immunity model.

More details can be found in the documentation and tutorials at `https://vg-sim.readthedocs.io/`)

### 7 Sampling

Sampled hosts are considered to be recovered and they are moved to a susceptible group (corresponding to the immunity triggered by the pathogen strain).

Sampling rates can be adjusted in each population to reflect differences in the sampling/sequencing efforts. Sampling rates can be also changed over time through the Python API between consequent calls of the `simulate()` method similarly to changing other parameters. It can be used, for example, to define the scenario where sampling begins with a delay after the pathogen first appears in the population.

We also note that it is straightforward for users to prune samples from the genealogy at any point in the simulation using matUtils [4] if this is thought to reflect a more realistic sampling scheme.

### 8 VGsim and other epidemiological simulators

Several epidemiological simulators were developed in recent years [5–13] to address many specific scenarios, each of which required certain trade-offs of general applicability vs realism and computational efficiency.

For example, agent-based simulators are able to tune models with extreme precision based on the exact properties of each agent. They also can model additional complex agent interactions — for example nosoi [8] can simulate complex geographical structures, while FAVITES [9] can model social contact networks. Accounting for geographical and social structure complexity increases the realism of the simulations, but this increased realism comes at a substantial cost in terms of computational efficiency. For example, in our tests, simulating a pandemic up to $10^7$ infected hosts with nosoi under a SIR-like model required more than a day.

SANTA-SIM is a software that can simulate complex scenarios of pathogen genome sequence evolution including selection, recombination, and indels [10]. While SANTA-SIM therefore employ a very general models of genome evolution, the choice of epidemiological models is not as broad as typical simulators that are specifically developed to simulate epidemiological dynamics.

PhyDyn [11] and TiPS [13] are also packages with simulation capabilities and similar forward-backward idea to genealogies. In [13] it was shown that TiPS is at least an order of magnitude faster than PhyDyn, so we did a more detailed comparison with the former.

Next we provide a more detailed comparison of VGsim with MASTER [14] and TiPS [13] simulators.

#### 8.1 MASTER

MASTER [14] is a general popular simulator which implements Gillespie algorithm, and it is built as a BEAST2 [15] package. It is used in many epidemiological studies [16–18]. In particular, in [19] MASTER was used for ABC inference, which suggests the potential use of VGsim for parameter estimation from real data.

MASTER does not have an option to sample individuals, instead it builds the full transmission chain of all the descendants of certain cases. This is a subset of VGsim's functionality, since the same behaviour can be achieved in our simulator by setting all recovery rates $\rho_h$ to zero and all the sampling rates $c^k\zeta_h$ to the intended recovery rates. With these settings, all recovered individuals are sampled and each isolate appears in the resulting tree. We ran the same SIR model in both simulators with transmission rate $\lambda = 2.7$, and recovery/sampling rate $\zeta = 1$. One can see that `VGsim` is considerably faster and scales better with the simulation size 1. For 10 million samples `MASTER` failed with an out of memory error on a machine with 16GB of memory. Moreover, by taking advantage of the fact that typically only a fraction of cases is sampled, and by efficiently sampling and storing only the genealogy of these samples instead of the whole epidemic transmission tree, `VGsim` allows to simulate large genealogies from massive pandemic scenarios.

`VGsim` also allows simulation scenarios not implemented in `MASTER`, such as the simulation of different haplotypes fitnesses and host immunity responses. Another new useful feature of `VGsim` is its intuitive and flexible interface. For example with 100 populations, 16 haplotypes and 3 immunity groups, there are approximately $5 \cdot 10^5$ reaction equations in the corresponding compartmental model.

`MASTER` has an option to approximate the simulated epidemiological dynamics with tau-leaping algorithm [20]. So, if the sample genealogy is not needed, `MASTER` should be the preferred way to simulate the epidemiological dynamics at least for simpler scenarios. For complicated models (especially with many populations), it is less clear if `VGsim` would be less effective compared to `MASTER` in simulating chains of events (only the forward simulation run) due to `VGsim`'s algorithm optimisations and caching of reaction rates. We did not yet explore this aspect in depth.

#### 8.2 TiPS

The forward-backward approach is also used in the `TiPS` package [13], which generates sampled trees based on epidemiological trajectories. It allows a flexible approach for generating sampled subtrees by giving the user substantial choice in terms of times of sampling. For example, sampling times might depend on models parameters — i.e. be proportional to the change in number of recovered individuals, similar to the approach in `VGsim`. For the simplest SIR model `VGsim` is about two times faster than `TiPS` with the direct Gillespie method (see Table 1). For the backward run, VGsim scales much better than `TiPS` (Table 2).

The focus of `VGsim` on complex epidemiological models is also captured in the software user interface, where `VGsim` allows a simplified specification of complex epidemiological dynamics.

Another important difference between the two simulators is the memory demand. We could not run `TiPS` for $3 \cdot 10^8$ iterations on our machines due to memory limitations, while on the same computers we could run `VGsim` for $2 \cdot 10^9$ iterations.

The strong point of `TiPS` is the implementation of hybrid and $\tau$-leaping algorithms which can reduce the computation time and memory demand by orders of magnitude. This is a feature that we would also like to include in `VGsim` in the future.

The primary purpose of `VGsim` is to generate sample genealogies from complex epidemiological models, and was developed with particular attention to its computational efficiency so to scale efficiently of global-scale pandemic datasets.

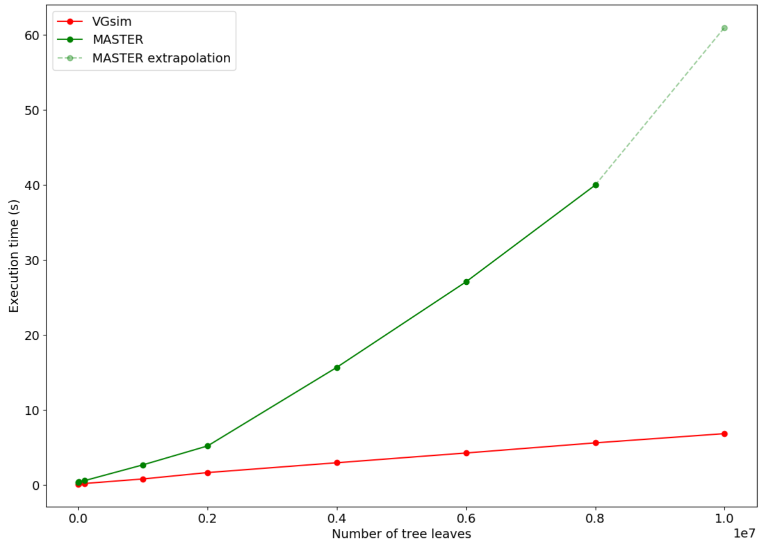


Figure 1: The comparison of `VGsim` and `MASTER` performance. Here we report the time to simulate a tree with a given number of leaves. `MASTER` crashed with an out of memory error for a tree with 10 million leaves, so we approximately extrapolated the final data point. The comparison was performed on a computer with Intel Core i7-7700K 4.20 GHz processor and 16GB of memory.

| Number of iterations | VGsim | TiPS |
|---|---|---|
| $10^6$ | 0.19 | 0.31 |
| $5 \cdot 10^6$ | 0.96 | 1.72 |
| $10^7$ | 1.84 | 3.44 |
| $5 \cdot 10^7$ | 9.87 | 17.57 |
| $10^8$ | 19.06 | 38.94 |

Table 1: Forward run time in seconds for different number of iterations under the SIR model. The recovery rate is set to 1 and the transmission rate to 2.5. The tests were run on a MacBookPro with Quad-Core Intel Core i5 2 GHz processor and 16GB of memory.

| Number of samples | VGsim | TiPS |
|---|---|---|
| $10^4$ | 0.059 | 242.4 |
| $2 \cdot 10^4$ | 0.11 | 808.2 |
| $3 \cdot 10^4$ | 0.18 | 1377.6 |
| $4 \cdot 10^4$ | 0.22 | 1921.2 |
| $5 \cdot 10^4$ | 0.27 | 3157.2 |

Table 2: Backward run time in seconds to generate genealogies for different sample sizes under the SIR model. The population size is $10^7$ in all runs. The recovery rate is 1 while the transmission rate is 2.5. The tests were run on a MacBookPro with Quad-Core Intel Core i5 2 GHz processor and 16GB of memory.

In addition, it can be used in other ways, for example, to generate random epidemiological trajectories, though there are ODE-based solutions such as `PhyDyn` that are better built for this specific purpose. To our knowledge, `VGsim` has a unique set of features when one needs to simulate an epidemiological model with a high level of complexity, including, but not limited to, the effects of selection, rare mutations, lockdowns, and migrations.
